# Independent Validation of Test-Adjusted COVID-19 Incidence Estimates Using Wastewater Surveillance Data in Ontario, Canada

**DOI:** 10.64898/2026.05.08.26352754

**Authors:** David N. Fisman, Natalie J. Wilson, Clara Eunyoung Lee, Ashleigh R. Tuite

## Abstract

**Background:** Case-based infectious disease surveillance is subject to ascertainment bias when testing intensity varies across time and population subgroups. We previously developed a regression-based test adjustment methodology using Standardized Testing Ratios (STRs) to correct for differential testing patterns in COVID-19 surveillance data. Wastewater-based surveillance (WWS) measures viral burden in the community independently of diagnostic testing behavior, making it a valuable external validation tool for test-adjusted case estimates.

**Methods:** We analyzed 111 weeks of paired wastewater and case surveillance data from Ontario, Canada (July 19, 2020 to August 28, 2022). Wastewater SARS-CoV-2 signals from 107 sewersheds across 34 public health units were normalized within sewersheds and aggregated using population-weighted averages. We compared wastewater correlations with crude reported and test-adjusted case counts using Spearman rank correlations, linear regression, and negative binomial distributed lag nonlinear models (DLNM), stratified by epidemic period.

**Results:** Test-adjusted cases correlated substantially more strongly with wastewater signals than crude reported cases overall (Spearman ρ = 0.849 vs. 0.679; linear R² = 0.609 vs. 0.191). The advantage of test adjustment was greatest during the Omicron wave, when population-level diagnostic testing contracted sharply following PCR eligibility restrictions (ρ = 0.924 vs. 0.604; R² = 0.815 vs. 0.470). DLNM incorporating the wastewater signal explained substantially more variance in test-adjusted than crude reported cases (McFadden pseudo-R² 0.898 vs. 0.776), despite similar lag-response structure for both outcomes.

**Conclusions:** Wastewater surveillance provides compelling independent validation of a previously described test adjustment methodology for COVID-19 case surveillance. The agreement between wastewater signals and test-adjusted cases was strongest precisely when testing scarcity was most severe, supporting the use of test adjustment to recover accurate infection dynamics from case surveillance data during periods of changing testing access and policy.

## INTRODUCTION

Case counting forms the backbone of public health surveillance, with identification and reporting of cases of disease mandated by public health statutes in many jurisdictions (1–4). The temporal patterns and geographic distribution of cases inform public health authorities about population risk and often drive health policy decisions (2, 3). However, case identification is rarely complete, and factors such as screening program implementation, changing test technologies, and variable testing intensity can substantially influence case counts and apparent disease incidence (3).

During the SARS-CoV-2 pandemic in Ontario, Canada, we observed that apparent changes in disease epidemiology over time were driven as much by changes in testing frequency as by true changes in infection patterns (5, 6). High-frequency testing in long-term care facilities, for example, made disease risk appear concentrated in older women, who constitute the majority of long-term care residents, despite evidence suggesting broader community transmission (6).

To enable fair comparisons across pandemic waves and strengthen understanding of disease epidemiology, we developed a regression-based methodology to adjust for testing frequency (5–7). This approach generates ‘test-adjusted’ incidence estimates that approximate what would have been observed had the entire population been tested with the same intensity as the most-tested groups. Key insights from this methodology included substantially higher infection incidence among young adults and adolescents than suggested by reported case counts(6), improved correlation between cases and subsequent mortality waves after test adjustment (5), and clearer associations between community mask mandates and infection reductions (7).

While previous validation efforts demonstrated internal consistency and biological plausibility, they shared potential limitations: mortality data came from the same surveillance system as case data, and policy evaluation analyses, though robust, relied on aggregate data susceptible to ecological fallacy. Independent validation using surveillance modalities that do not share case surveillance biases is therefore crucial.

Wastewater surveillance emerged during the pandemic as a complementary approach to traditional case-based surveillance (8–11). Unlike case detection, which depends on individuals seeking testing and healthcare access, wastewater monitoring captures viral shedding from all infected individuals within a sewershed, regardless of symptoms, testing behavior, or healthcare engagement (8). This makes wastewater surveillance particularly valuable as an independent validation tool: it measures the same underlying phenomenon (population infection burden) through a completely different mechanism that is blind to testing patterns.

The Ontario COVID-19 Science Table participated in a comprehensive SARS-CoV-2 wastewater surveillance effort led by Ontario’s Ministry of Environment, Conservation and Parks. The effort was supported by the Canadian National Microbiology as well as a number of Canadian research groups, and continued across Ontario throughout much of the pandemic, providing an opportunity for rigorous external validation (12). Our objective was to validate the test-adjustment methodology by assessing whether test-adjusted incidence estimates were more strongly correlated with wastewater viral burden than unadjusted case counts.

## METHODS

### Study Period, Case Counts and Test-adjustment

We analyzed weekly Ontario COVID-19 case, testing, and wastewater surveillance data from July 19, 2020 to August 28, 2022, corresponding to the period in which wastewater and case surveillance data overlapped. Wastewater surveillance began on July 19, 2020 and expanded progressively over the study period. Weekly reported COVID-19 case counts and testing volumes for Ontario were obtained from the provincial Case and Contact Management System (CCM) as reported elsewhere (5, 6). Test adjustment methodology has been described in detail previously (5, 6). Briefly, we used indirect standardization with meta-regression to estimate expected case counts for each age-sex stratum based on observed testing intensity, expressed as Standardized Testing Ratios (STRs). Standardized Infection Ratios (SIRs) were then calculated as the ratio of observed to expected cases within each stratum. Test-adjusted case counts were derived by applying these SIRs to estimate the case counts that would have been observed had all strata been tested at the intensity of the most-tested reference group (women aged 80 years and older).

### Wastewater Surveillance Data

Wastewater data were obtained from provincial data files made available by the Ontario COVID-19 Science Table, based on measurements from wastewater treatment facilities across Ontario sewersheds (12). The first measurements began in Hamilton, Ontario in mid-July 2020, with additional sites added progressively throughout the study period. By the end of the study period, surveillance covered 107 wastewater treatment facilities serving approximately 11.8 million people (79% of Ontario’s population) (**Figure 1** and **Figure 2**).

**Figure 1.**
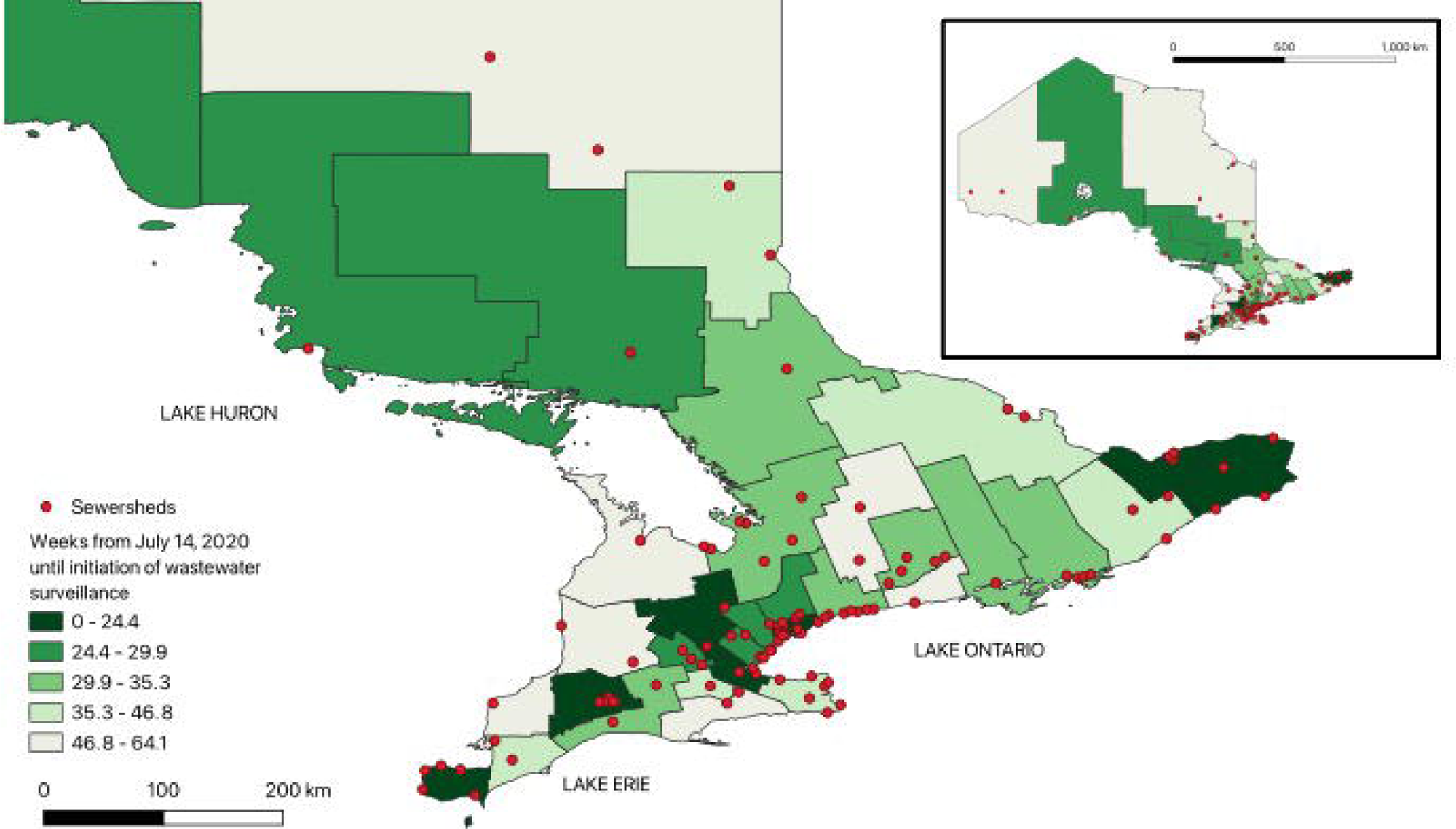
Geographic distribution of wastewater surveillance sewersheds across Ontario public health units, July 2020–August 2022. Ontario public health units (PHUs) are shaded according to the number of weeks of wastewater-based surveillance data contributed to the analysis, from earliest adopters (darkest green) to latest adopters (lightest green). Red circles indicate individual sewershed sampling locations. The main map features southern Ontario health units; the inset shows the entire province including northern Ontario PHUs at reduced scale. A total of 107 sewersheds across all 34 Ontario PHUs contributed data over the study period.

**Figure 2.**
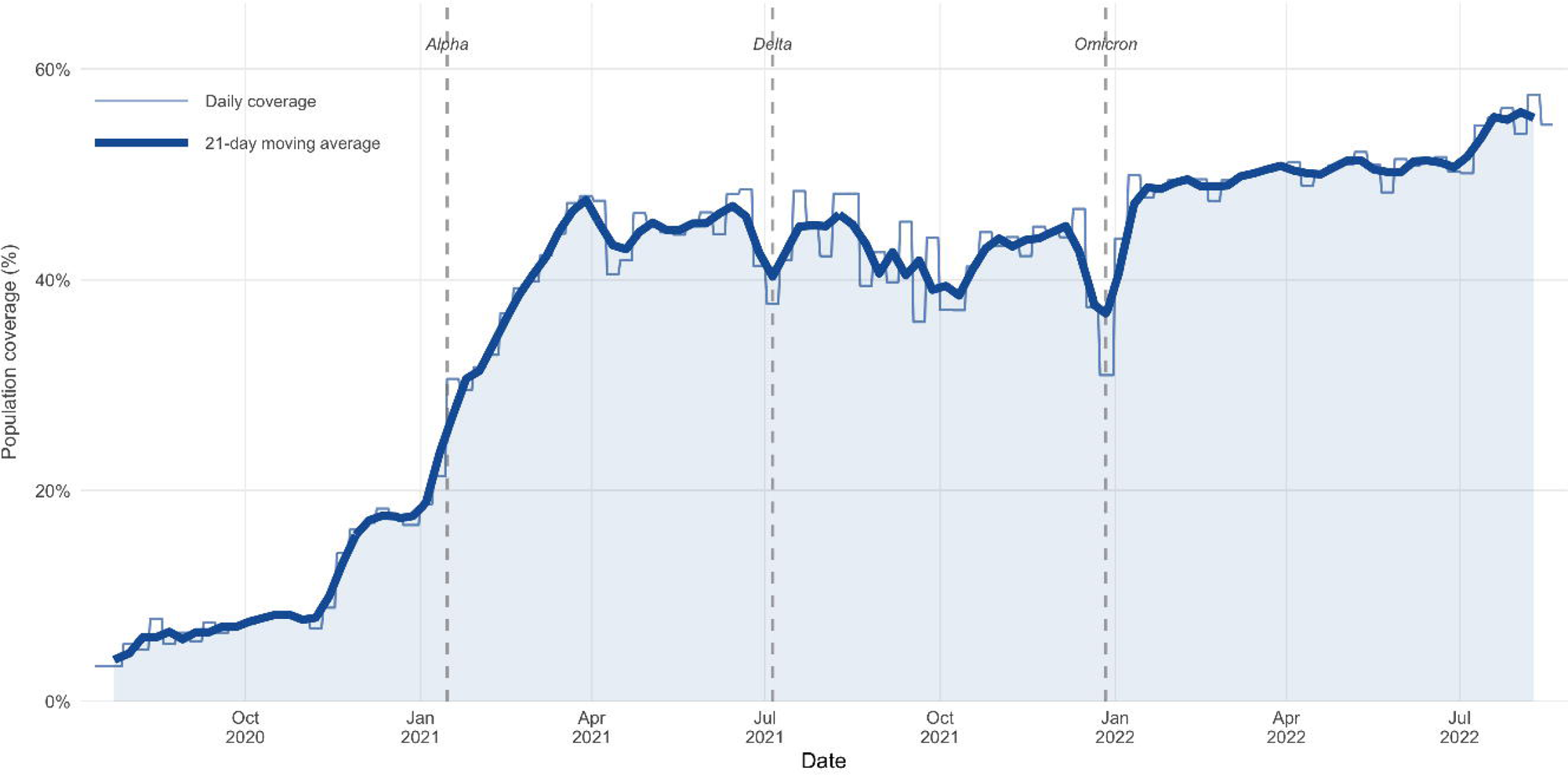
Proportion of the Ontario population served by participating wastewater treatment facilities, July 2020–August 2022. Population coverage was calculated as the sum of populations served by contributing sewersheds divided by the total Ontario population. The faint line shows weekly values; the bold line shows the 21-day centred moving average. Dashed vertical lines indicate the approximate onset of the Alpha (January 2021), Delta (July 2021), and Omicron (December 2021) variant waves in Ontario. Coverage expanded from approximately 3% at study initiation to a peak of 58% in August 2022, with the majority of scale-up occurring during 2021.

SARS-CoV-2 burden was quantified through measurement of N1 and N2 gene targets (11, 12). Because measurement protocols varied between sewersheds with respect to flow rate, sampling frequency, and analytical methods, we created a common metric by normalizing gene detection values within each sewershed by dividing by the standard deviation of its full time series. When both N1 and N2 targets were detected on the same date at the same location, we used their mean as the normalized signal; when only one target was detected, that value was used.

To estimate a province-wide signal, we calculated population-weighted averages for each day by multiplying each sewershed’s normalized signal by its served population, summing across all sewersheds with available data, and dividing by the total population served by contributing sewersheds on that day. Daily values were then averaged within each calendar week to produce the weekly wastewater signal used in all analyses.

### Statistical Analysis

We merged wastewater and case data by matching case reporting weeks to overlapping wastewater measurement periods, yielding 111 weeks of paired observations from July 19, 2020 through August 28, 2022. For all analyses, wastewater signal values were the within-sewershed SD-normalized, population-weighted weekly means described above; case counts (both reported and test-adjusted) were used on their original (non-normalized) scale.

We evaluated the strength of association between the wastewater signal and each case series using Spearman rank correlation coefficients. We also fit ordinary least squares regression models on both the original and log-transformed scales to obtain coefficients of determination (R²). Analyses were stratified by epidemic period (pre-Omicron and Omicron) to evaluate whether the advantage of test adjustment varied with testing intensity.

To characterize the temporal structure of the wastewater–case relationship while allowing for nonlinearity and distributed lag effects, we fit distributed lag nonlinear models (DLNM) using a negative binomial generalized linear model framework. A cross-basis matrix was constructed for the wastewater signal using natural cubic splines with 3 degrees of freedom in both the exposure and lag dimensions, with a maximum lag of 8 weeks and centering at zero (no wastewater signal). A natural cubic spline with 2 degrees of freedom for calendar time was included to account for secular trends. Separate models were fit using reported and test-adjusted cases as the outcome. We report lag-specific exposure–response associations expressed as rate ratios, along with 95% confidence intervals. Model fit was summarized using the McFadden pseudo-R², and the incremental contribution of the wastewater signal was assessed by comparing full models against trend-only null models using likelihood ratio tests.

All analyses were performed in R version 4.5.0 (R Core Team, 2025). Key packages included dlnm (v2.4.2) for distributed lag nonlinear models, MASS for negative binomial regression, and ggplot2 and patchwork for figure generation. Analysis code and data are available at https://github.com/fismanda/wastewater; DOI: 10.5281/zenodo.19373810. The study was approved by the Research Ethics Board of the University of Toronto (protocol #41690).

## RESULTS

### Wastewater Surveillance Coverage

Wastewater-based surveillance data were contributed by 107 sewersheds across all 34 Ontario public health units (PHUs) over the study period. Coverage expanded substantially over time, from approximately 3% of the Ontario population at study initiation in July 2020 to a peak of 58% in August 2022 (**Figures 1** and **2**). In the earliest weeks of the study period, surveillance was limited to two PHUs (Hamilton and Ottawa), with broader provincial coverage achieved progressively over 2021. The geographic distribution of sewershed locations and the timeline of coverage expansion are shown in **Figures 1** and **2**.

### Descriptive Statistics

Over the 111-week study period, mean weekly reported cases were 12,072 (SD 15,714; range 565–107,648), while mean test-adjusted cases were 41,392 (SD 46,976; range 1,374–238,146). Test adjustment increased cumulative case estimates by 243% over the study period. Mean normalized wastewater signal was 0.54 (SD 0.51; range 0.01–2.48). **Figure 3** illustrates the temporal relationship between the normalized wastewater signal and COVID-19 case counts over the study period. Crude reported cases and test-adjusted case estimates tracked closely together, and both tracked the wastewater signal, during periods of high testing intensity including the Alpha and Delta waves of 2021. A divergence emerged during the Omicron wave beginning in late December 2021, when population-level diagnostic testing contracted sharply (5). During this period, the wastewater signal and test-adjusted cases both indicated a sharp, large-magnitude epidemic peak, while crude reported cases remained substantially attenuated. Peak test-adjusted cases during the week of January 4, 2022 were approximately 2.3-fold higher than peak crude reported cases (211,693 vs. 93,768); the ratio of adjusted to reported cases reached 3.7-fold the following week and continued to rise throughout 2022, exceeding 11-fold by August 2022 as population-level testing wound down (**Figure 3**).

**Figure 3.**
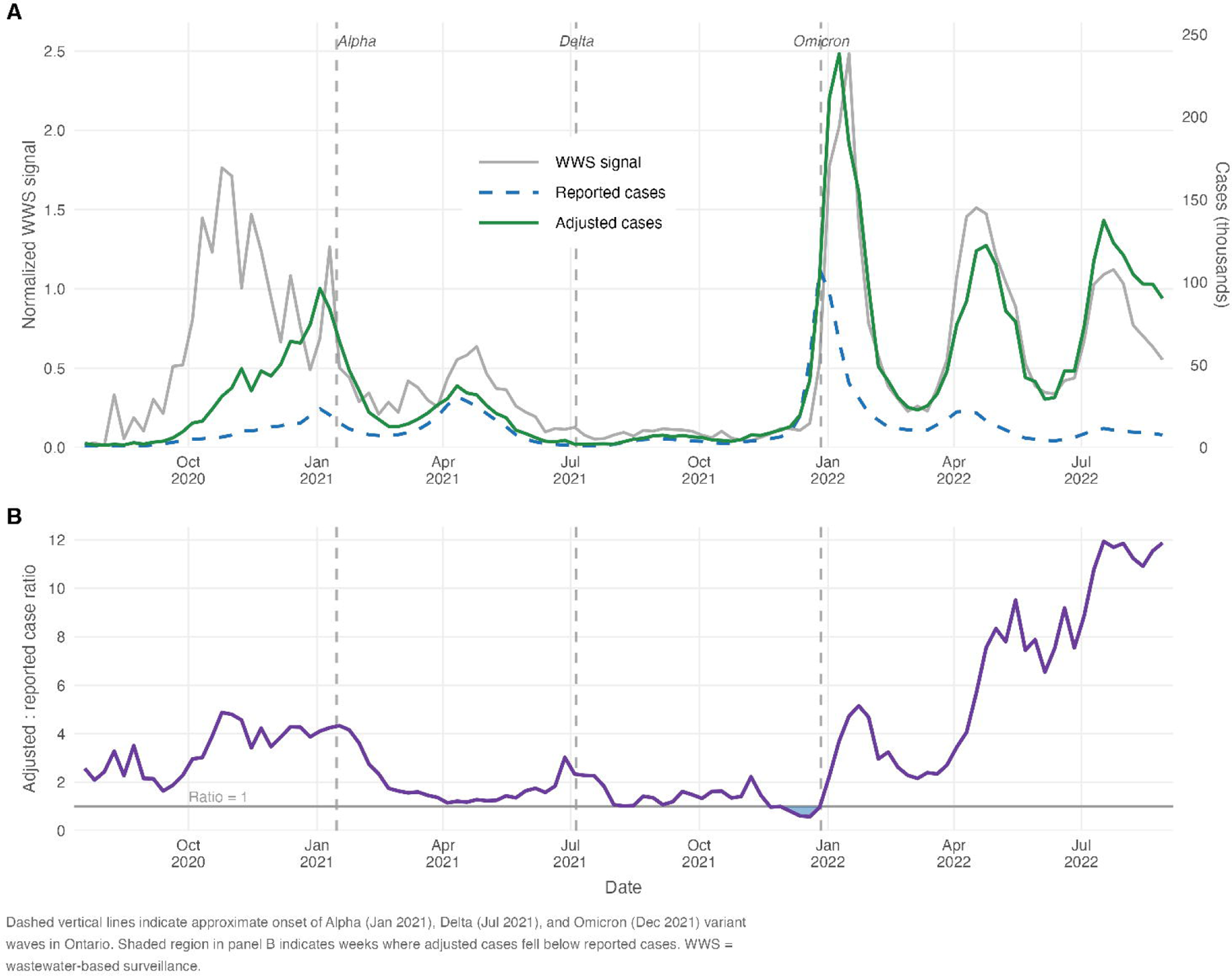
Temporal relationship between wastewater-based SARS-CoV-2 surveillance signal, crude reported COVID-19 cases, and test-adjusted case estimates, Ontario, Canada, July 2020–August 2022. (A) Weekly normalized wastewater signal (left axis, grey) plotted alongside weekly crude reported cases (right axis, blue dashed) and test-adjusted case estimates (right axis, green) over the 111-week study period. The wastewater signal was normalized by standard deviation within each sewershed prior to population-weighted aggregation across 107 sewersheds serving 34 Ontario public health units. (B) Ratio of test-adjusted to crude reported cases over time. The horizontal line indicates a ratio of 1.0 (adjusted equals reported). Shaded region indicates weeks in which adjusted cases fell below reported cases. Dashed vertical lines indicate the approximate onset of the Alpha (January 2021), Delta (July 2021), and Omicron (December 2021) variant waves in Ontario. The marked divergence between panels beginning in late December 2021 reflects the sharp contraction of population-level diagnostic testing during the Omicron wave. WWS, wastewater-based surveillance.

### Correlation and Linear Regression Analysis

Wastewater-based surveillance signals were more strongly correlated with test-adjusted than with crude reported COVID-19 cases. Spearman rank correlation between the normalized wastewater signal and test-adjusted cases was ρ = 0.849 (p < 10□^3^□), compared with ρ = 0.679 (p < 10□^1^□) for crude reported cases, representing a 25% relative improvement in correlation attributable to test adjustment. Linear regression on the case count scale explained 60.9% of variance in test-adjusted cases (R^2^ = 0.609) compared with 19.1% for crude reported cases (R² = 0.191), a 3.2-fold difference. When we regressed log-transformed cases on the wastewater signal, this specification yielded R^2^ = 0.722 for test-adjusted and R² = 0.464 for crude reported cases. Scatter plots for all four regression models are shown in Figure 4.

**Figure 4.**
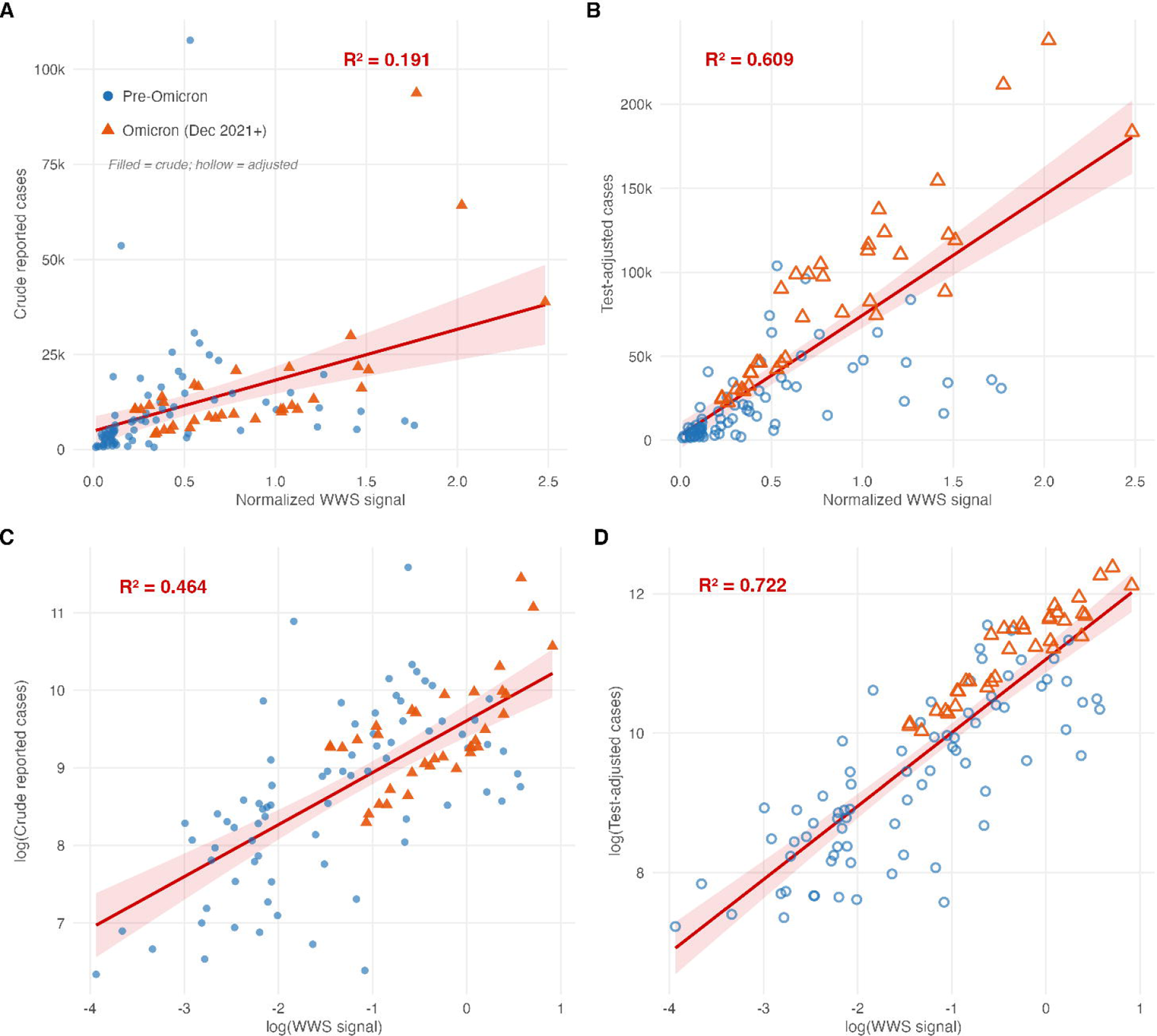
Association between normalized wastewater-based surveillance signal and COVID-19 case counts, Ontario, Canada, July 2020–August 2022. Association between normalized wastewater-based surveillance (WWS) signal and COVID-19 case counts, Ontario, July 2020 to August 2022. Panels A and B show linear-scale regression of crude reported and test-adjusted cases on the WWS signal, respectively. Panels C and D show the corresponding log-log regressions. Filled symbols indicate crude reported cases (A, C); hollow symbols indicate test-adjusted cases (B, D). Circles denote pre-Omicron weeks; triangles denote Omicron-period weeks (December 27, 2021 onward). Red lines indicate ordinary least squares regression fits with 95% confidence bands (shaded). R² values are shown for each panel.

Stratification by epidemic period revealed that the advantage of test-adjusted over crude reported cases was most pronounced during the Omicron wave (**Table 1**). During the Omicron period, Spearman correlation between the wastewater signal and test-adjusted cases was ρ = 0.924 (95% CI 0.853–0.961), compared with ρ = 0.604 (95% CI 0.340–0.781) for crude reported cases; linear R² was 0.815 versus 0.470 respectively. In the pre-Omicron period, correlations were more similar between the two series (ρ = 0.786 vs. 0.669; R² = 0.323 vs. 0.037), though the low linear R² for crude reported cases in this period likely reflects both the limited geographic coverage of the wastewater network in early weeks of the study (restricted to Hamilton and Ottawa) and variable test-seeking behaviour during a period when testing intensity tracked disease activity rather than systematic surveillance policy.

### Distributed Lag Nonlinear Models

Negative binomial DLNMs incorporating the wastewater cross-basis substantially improved model fit over trend-only null models for both crude reported and test-adjusted cases. The incremental improvement in McFadden pseudo-R² attributable to the wastewater signal was 0.619 for reported cases (trend-only: 0.157; full model: 0.776) and 0.683 for adjusted cases (trend-only: 0.214; full model: 0.898), with both likelihood ratio tests significant (p < 0.001). The lag-response curves at the 75th percentile of the wastewater signal were similar in shape for both outcomes, with the strongest association at lag 0 declining monotonically through lag 8 weeks (**Figure 5**). The superior overall model fit for test-adjusted cases, despite near-identical lag structure, suggests that test-adjusted cases track the wastewater signal more closely than crude reported cases. Lagged Spearman correlations were consistently higher for test-adjusted than crude reported cases across all lags from 0 to 8 weeks (**Figure 5**), with correlations of ρ = 0.849 and 0.679 respectively at lag 0, declining to ρ = 0.296 and 0.152 at lag 8 weeks.

**Figure 5.**
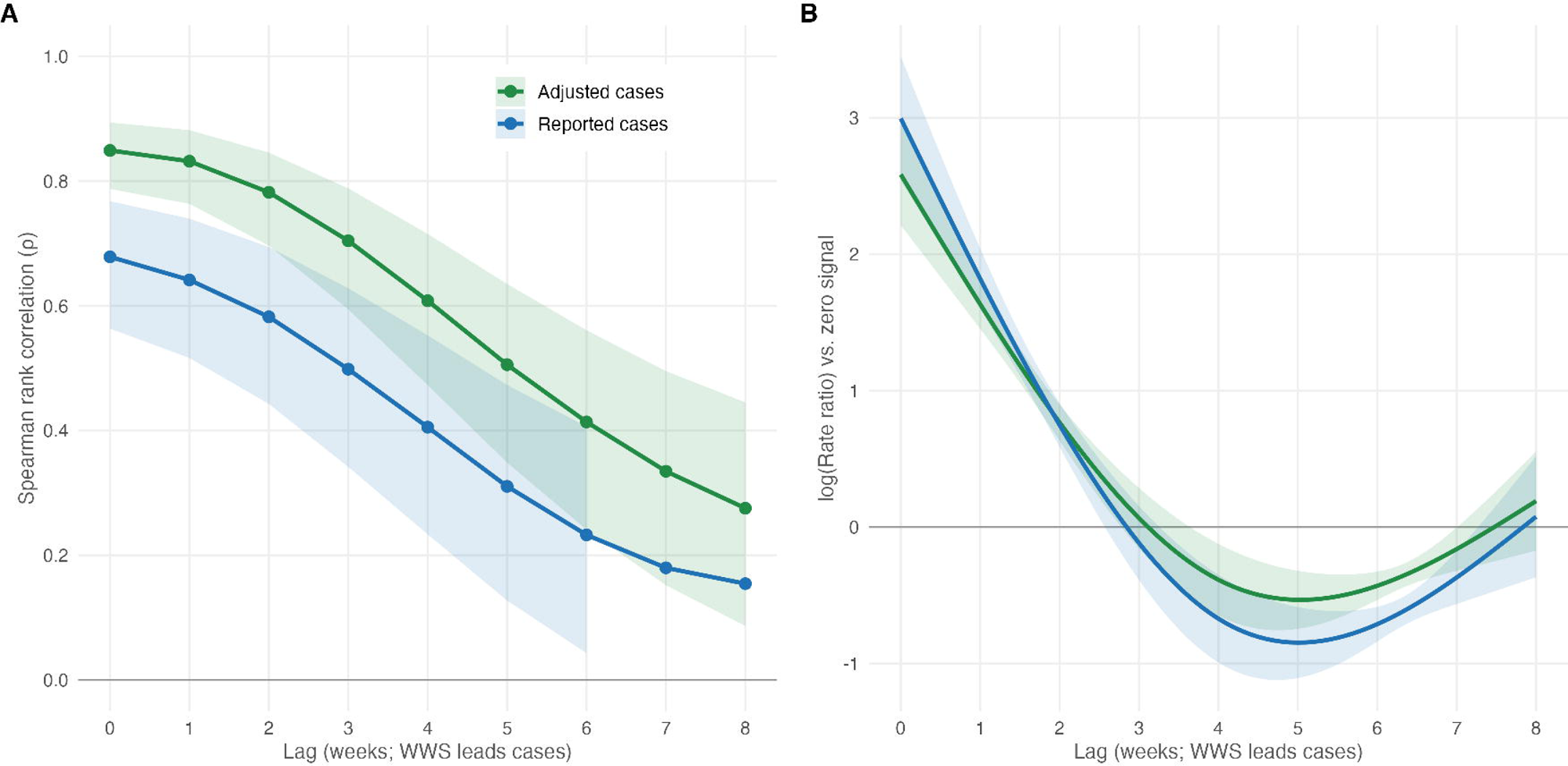
Temporal association between normalized wastewater-based surveillance signal and COVID-19 case counts across lags of 0 to 8 weeks, Ontario, Canada, July 2020–August 2022. (A) Spearman rank correlation between the normalized wastewater signal at time *t* and weekly case counts *k* weeks later, for *k* = 0 to 8 weeks. Correlations are shown separately for crude reported cases (blue) and test-adjusted cases (green). Shaded bands indicate 95% confidence intervals derived via Fisher z-transformation. (B) Lag-response curves from negative binomial distributed lag nonlinear models (DLNM), showing the log rate ratio for weekly case counts relative to a zero wastewater signal, evaluated at the 75th percentile of the normalized wastewater signal (WWS = 0.767). Models included a cross-basis matrix with natural splines (3 degrees of freedom) in both the exposure and lag dimensions, a maximum lag of 8 weeks, and a natural spline trend term (2 degrees of freedom). Shaded bands indicate 95% confidence intervals. The reference line at log(RR) = 0 corresponds to a rate ratio of 1.0 (null association). The dip below zero at lags 4–5 weeks reflects spline effects rather than a true negative association.

## DISCUSSION

In this study, test-adjusted COVID-19 case estimates aligned more closely with Ontario’s wastewater SARS-CoV-2 signal than did crude reported cases across multiple analytic approaches, particularly during the Omicron period. Wastewater surveillance provides a useful external benchmark for evaluating this question because it is not influenced by healthcare seeking and diagnostic test access in the same way as case-based surveillance. Previous work showed that test-adjusted cases aligned more closely than crude reported cases with other downstream indicators of epidemic activity, including mortality and critical care burden (5, 13). The present findings extend that validation to an independent surveillance stream.

The advantage of test adjustment was most pronounced during the Omicron wave, when population-level diagnostic testing contracted sharply following Ontario’s restriction of publicly funded PCR testing to narrower high-risk groups on December 30, 2021 (14). During this period, the wastewater signal remained closely aligned with test-adjusted cases, while crude reported cases appeared substantially attenuated. Peak test-adjusted cases in the week of January 4, 2022 were 2.3-fold higher than peak crude reported cases, and the adjusted-to-reported ratio continued to widen through 2022 as population-level testing wound down. This pattern is consistent with the interpretation that crude reported cases became increasingly shaped by testing policy and behaviour, whereas the adjusted series better preserved the underlying epidemic signal.

In the pre-Omicron period, the gap between adjusted and crude reported cases was smaller, consistent with a time when broader PCR testing access and higher overall testing intensity made reported cases a more stable proxy for transmission trends. The relatively weak linear fit for crude reported cases in this period likely reflects both the limited geographic coverage of the wastewater network in the earliest weeks of the study and variable test-seeking behaviour during a period when testing intensity tracked disease activity rather than a systematic surveillance policy. Even so, test-adjusted cases remained more strongly associated with the wastewater signal, suggesting that adjustment partially compensates for non-systematic testing patterns.

Lagged analyses further support the interpretation that wastewater and case series tracked the same epidemic waves at the temporal resolution available here. The strongest associations were observed contemporaneously, with progressively weaker associations at longer lags, and distributed lag nonlinear models showed better overall fit for test-adjusted than crude reported cases. The similar shape of the lag-response curves for the two outcomes suggests that both series reflected the same broad temporal dynamics, while the stronger fit for adjusted cases indicates closer agreement with the wastewater signal. The predominantly contemporaneous association is also consistent with the use of weekly aggregation: lead times of only a few days between viral shedding and clinical detection may be detectable at daily resolution, but are likely compressed within the same weekly measurement window.

Wastewater surveillance is not a perfect gold standard for true incidence. Fecal shedding of SARS-CoV-2 RNA occurs in a substantial fraction of infected individuals, including those with asymptomatic infection, but shedding magnitude and duration vary across individuals, viral lineages, and time (15). For this reason, wastewater is best viewed as a population-level trend indicator rather than a precise measure of incidence. Even so, its value as a benchmark is strengthened by prior work showing that SARS-CoV-2 wastewater concentrations predict COVID-19 hospital admissions across multiple Canadian cities and variant periods (15). Together with previous findings for mortality and critical care admissions, the present results add to a consistent body of evidence that test-adjusted cases align more closely than crude reported cases with independent indicators of epidemic activity.

These findings have important implications for public health surveillance. Testing patterns changed substantially throughout the COVID-19 pandemic because of shifting eligibility criteria, resource constraints, and public behaviour. As a result, reported case counts did not always provide a stable or comparable picture of infection dynamics across waves. Test adjustment offers one way to improve the interpretability of case-based surveillance when testing access and policy change, and may help preserve comparability across periods in which crude case counts are especially vulnerable to distortion.

The potential relevance of this approach extends beyond COVID-19. Many infectious diseases are subject to differential testing across populations because of access barriers, variation in clinical presentation, or targeted surveillance programs. In those settings, methods that explicitly account for observed testing patterns may improve inference about disease burden from routine surveillance data. We have previously applied this framework to pertussis, where testing intensity also varies substantially across age groups (16), suggesting that this type of adjustment may be useful in other surveillance contexts as well.

Several limitations should be considered. Wastewater measurements varied across sewersheds in sampling protocols, flow and dilution characteristics, and laboratory methods; although our normalization approach reduces some of this heterogeneity, residual differences likely remain. The wastewater surveillance network also expanded substantially over the study period, introducing time-varying heterogeneity in geographic coverage that population-weighted aggregation can only partially address. Results from the earliest weeks of the study should therefore be interpreted cautiously. Finally, weekly aggregation may obscure finer temporal relationships between wastewater and case detection, and daily-resolution analyses would be better suited to evaluating short biological lead times.

Overall, Ontario wastewater surveillance provided an independent benchmark against which test-adjusted COVID-19 case estimates aligned more closely than crude reported cases, particularly during periods of constrained diagnostic testing. These findings support the usefulness of test adjustment for improving interpretation of case-based surveillance when testing access and policy change over time.

## Data Availability

Analysis code and data are available at https://github.com/fismanda/wastewater; DOI: 10.5281/zenodo.19373810.

## ACKNOWLEDGEMENTS

We thank the Ontario COVID-19 Science Table for provision of wastewater surveillance data. This work was supported by the Canadian Institutes for Health Research (OV4-170360 and #518192) to Dr. Fisman.

